# First-week expansion of activated peripheral B-cell subsets is associated with grade 3 or higher immune-related adverse events after nivolumab plus ipilimumab

**DOI:** 10.64898/2026.09.09.26362435

**Authors:** Hidetomo Takakura, Yohei Funakoshi, Taiji Koyama, Tomoki Sasaki, Eriko Fukuda, Jae Man Lee, Keiji Kurata, Yumiko Inui, Kimikazu Yakushijin, Shiro Kimbara, Yoshiaki Nagatani, Naomi Kiyota, Hironobu Minami

**Affiliations:** Division of Medical Oncology/Hematology, Department of Medicine, Kobe University Hospital and Graduate School of Medicine, Kobe, Japan; R&D Department, KAICO Ltd., Fukuoka, Japan; Cellular and Molecular Biotechnology Research Institute, National Institute of Advanced Industrial Science and Technology (AIST), Ibaraki, Japan; Laboratory of Insect Food Sciences, Kyushu University Graduate School of Bioresource and Bioenvironmental Sciences, Fukuoka, Japan; Cancer Center, Kobe University Hospital, Kobe, Japan

**Keywords:** immune checkpoint inhibitors, immune-related adverse events, B cells, nivolumab, ipilimumab, B-cell receptor repertoire, myasthenia gravis

## Abstract

**Background:** Although immune checkpoint inhibitors (ICIs) cause grade 3 or higher immune-related adverse events (irAEs), validated biomarkers for identifying patients at risk of these events are lacking. Given the potential contribution of B-cells to irAE development, we evaluated whether early changes in circulating B-cell subsets after nivolumab plus ipilimumab were associated with subsequent severe irAEs.

**Materials and Methods:** This prospective observational study included 12 patients who received nivolumab plus ipilimumab, and an exploratory cohort of five patients who received anti-PD-1 monotherapy. Blood was sampled before treatment, at week 1, and before cycle 2. Flow cytometry quantified CD21^low^ B-cells, CD27^+^IgD^−^ class-switched B-cells, plasmablasts, and CD19^+^CD138^+^ plasma cells. irAEs were assessed for 6 months, and fold changes at week 1 were compared between patients with and without severe irAEs. Paired immunoglobulin G B-cell receptor (BCR) repertoires were analyzed in four patients with grade 3 or higher irAEs. One patient developed myasthenia gravis, and anti-acetylcholine receptor (AChR) antibody was measured in serial serum samples.

**Results:** In the nivolumab plus ipilimumab patients, grade 3 or higher irAEs occurred in five patients. Week-1 increases in all four activated B-cell subsets were significantly greater in these patients, whereas total CD19^+^ B-cell changes did not differ. Rarefied IgG BCR diversity generally increased in repertoires derived from peripheral blood mononuclear cells, while Gini coefficients decreased in all paired repertoires. In the myasthenia gravis case, anti-AChR antibody exceeded the reference range by Day 11, before symptoms and diagnosis.

**Conclusion:** First-week expansion of activated peripheral B-cell subsets after nivolumab plus ipilimumab was associated with subsequent severe irAEs. These findings support future studies of early B-cell monitoring within integrated multimarker assessments of irAE risk.

## 1 Introduction

Immune checkpoint inhibitors (ICIs) targeting programmed cell death protein 1 (PD-1) and cytotoxic T-lymphocyte antigen 4 (CTLA-4) restore antitumor immune activity by blocking inhibitory pathways that maintain immune tolerance (1, 2). In recent clinical trials, combined PD-1 and CTLA-4 blockade was effective in melanoma, renal cell carcinoma, and other cancers (3, 4). Although ICI therapy has the potential to activate immune cells against tumor cells in patients, it can also cause specific toxicities known as immune-related adverse events (irAEs). Examples include dermatitis, colitis, hepatitis, thyroid dysfunction, adrenal insufficiency and myasthenia gravis (5). In pivotal trials of renal cell carcinoma and melanoma, grade 3 or higher treatment-related adverse events occurred in 46%–59% of patients receiving nivolumab plus ipilimumab and were more frequent than in anti-PD-1 monotherapy (3, 4). Severe irAEs may be life-threatening or fatal and may require treatment discontinuation, systemic immunosuppression, hospitalization, or specialist care (5).

Therefore, early diagnosis is essential. To date, however, no validated biomarker for patients at risk of irAEs has been identified (6, 7).

Immune checkpoint molecules are receptors that are generally expressed by immune cells and are particularly relevant to T-cell activity. CD8^+^ T-cell infiltration has been reported in organs affected by irAEs after PD-1 blockade, alone or with CTLA-4 blockade (8, 9). Although checkpoint molecules are most closely associated with T-cell regulation, they also contribute to B-cell tolerance (10). Because some irAEs share features with autoimmune diseases and can involve autoantibodies, B-cell populations may be relevant to irAE development (11). In their study of patients with renal cell carcinoma receiving nivolumab plus ipilimumab, Uehara et al. reported that the differentiation of circulating B-cells after treatment was enhanced among patients who developed severe irAEs, and included an increase in plasmablasts (12). Moreover, Gatto et al. linked irAE-associated arthritis to expansion of transitional CD10^+^CD24^high^CD38^high^ B-cells (13). These findings suggest that changes in activated or differentiating B-cell populations may be associated with irAE development. However, the very early time course of B-cell changes after ICI administration and their relationship to subsequent severe irAEs remain unclear.

irAEs most commonly develop within 2–3 months after ICI initiation but may occur at any time during treatment or even much later thereafter (14, 15). The timing of immune activation leading to irAEs after ICI initiation has not been established (6, 14). Importantly, changes in the immune system in target organs are already occurring even in the asymptomatic stage, before clinical symptoms are recognized (13, 16, 17). These observations suggest that immunologic changes relevant to irAE development may precede their clinical manifestations. Accordingly, detailed characterization of the immune state during the early phase after ICI initiation may capture immunologic changes before irAEs become clinically apparent.

Based on these considerations, we hypothesized that early changes in activated B-cell subsets after initiation of ICI would be associated with the subsequent development of severe irAEs. Here, to address this hypothesis, we characterized early peripheral B-cell changes in detail across multiple activated B-cell subsets after nivolumab plus ipilimumab and examined their association with subsequent grade 3 or higher irAEs.

## 2 Materials and Methods

### 2.1 Study design and patients

In this study, patients with malignant diseases who received ICI therapy, including nivolumab plus ipilimumab combination therapy or anti-PD-1 antibody monotherapy, at Kobe University Hospital between May 2019 and September 2020 were enrolled. Twelve patients received nivolumab plus ipilimumab, and five received anti-PD-1 monotherapy.

Clinical information was obtained from medical records and routine hospital visits. irAEs were graded according to the National Cancer Institute Common Terminology Criteria for Adverse Events, version 5.0. In this study, irAEs of grade 3 or higher were defined as severe irAEs. Clinical outcome was the occurrence of at least one severe irAE within 6 months after the first ICI initiation.

The study was approved by the Kobe University Hospital Ethics Committee (approval numbers B2056704 and 1481) and was conducted in accordance with the Declaration of Helsinki. All patients provided written informed consent for participation and sample collection.

### 2.2 Blood sampling and sample processing

Blood sampling was performed pretreatment (within 2 days before the first dose), 1 week after the first dose (Day 7 ± 1), and before cycle 2 (within 2 days before the next dose) (Figure 1). In two patients, additional blood samples were collected during the first cycle or after subsequent cycles, as outlined in the clinical study protocol.

**Figure 1.**
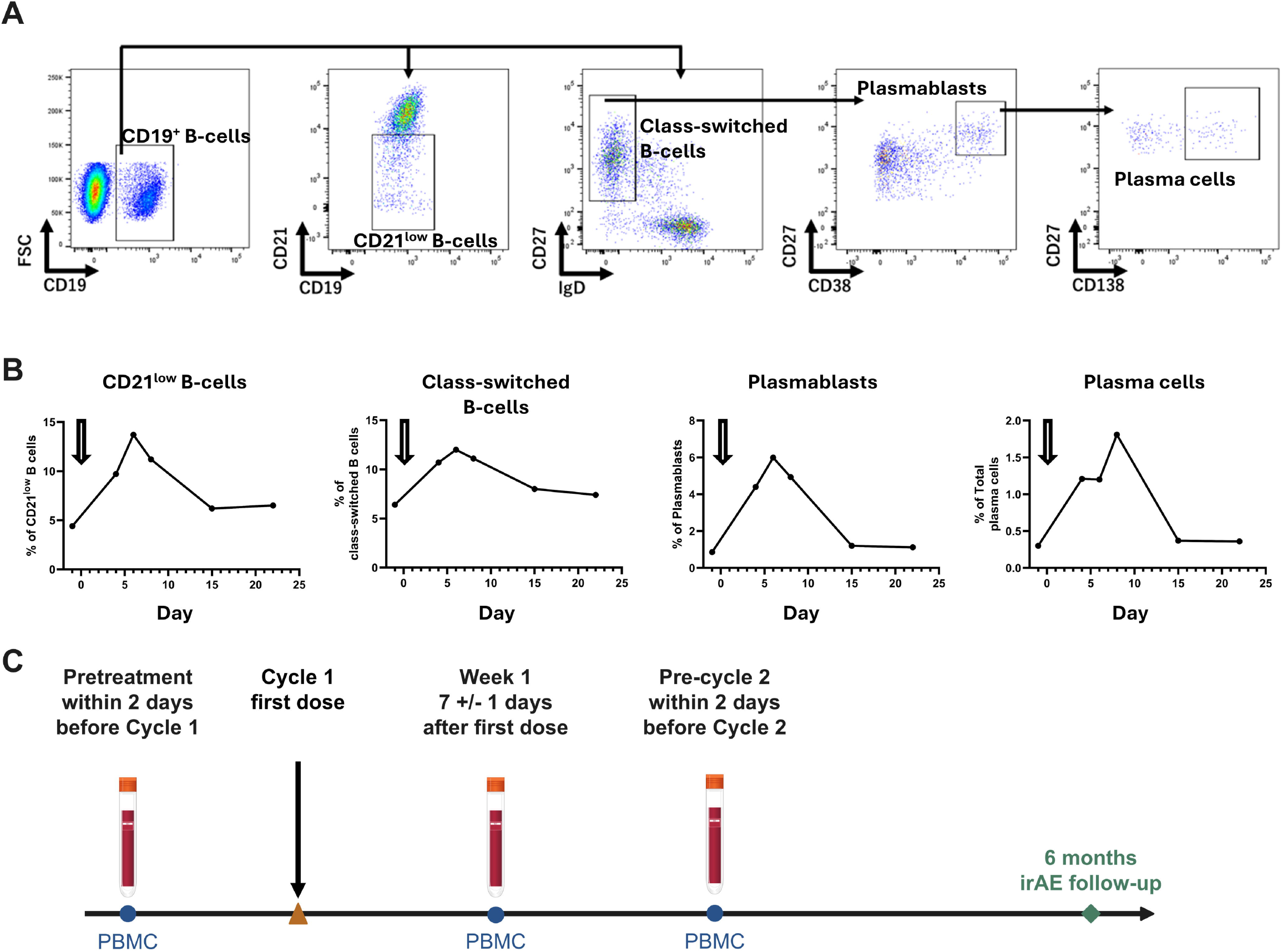
Study design, serial blood sampling, and flow-cytometry gating strategy. (A) Representative flow-cytometry gating strategy. After gating on broad mononuclear cells encompassing the lymphocyte and monocyte regions, CD19^+^ B-cells were identified. CD21^low^ B-cells and CD27^+^IgD^−^ class-switched B-cells were defined within the CD19^+^ B-cell gate. Plasmablasts were defined as CD27^high^CD38^high^ cells within the class-switched B-cell gate. CD19^+^CD138^+^ plasma cells were defined as the CD138^+^ subset within the plasmablast gate. (B) Serial changes in circulating B-cell populations in a patient evaluated separately in a hypothesis-generating analysis. These changes informed selection of the week-1 assessment for the 12-patient cohort analysis. B-cell subset frequencies were expressed as percentages of CD19^+^ B-cells. The downward arrows indicate the first administration of nivolumab plus ipilimumab. (C) Blood-sampling schedule. PBMC samples were collected within 2 days before cycle 1, at week 1 after the first administration (Day 7 ± 1), and within 2 days before cycle 2. irAEs were assessed for 6 months after ICI initiation. irAE, immune-related adverse event; PBMC, peripheral blood mononuclear cell. Alt text: Representative flow-cytometry plots show sequential identification of CD19^+^ B-cells, CD21^low^ B-cells, class-switched B-cells, plasmablasts, and CD138^+^ plasma cells. Serial measurements in a hypothesis-generating patient show transient increases in activated B-cell subsets approximately 1 week after treatment. The study schedule shows blood sampling before treatment, at week 1, and before cycle 2, with immune-related adverse events assessed for 6 months.

Peripheral blood was collected in heparinized tubes. Peripheral blood mononuclear cells (PBMCs) were isolated by density-gradient centrifugation using Ficoll-Paque Plus (Cytiva, Uppsala, Sweden) and SepMate-50 (STEMCELL Technologies, Vancouver, BC, Canada) tubes according to the manufacturers’ instructions. PBMCs were cryopreserved in CELLBANKER (Nippon Zenyaku Kogyo Co., Ltd., Koriyama, Fukushima, Japan) and stored at −80°C until analysis. Serum was separated by centrifugation for 10 minutes at 1,000 × g at room temperature and stored at −80°C.

### 2.3 Flow cytometry

PBMCs were stained for 20 minutes at 4°C with anti-human CD19 BV510, CD27 BV421, IgD FITC, CD38 BV711, and CD138 APC antibodies (BioLegend, San Diego, CA, USA), together with CD21 PE and 7-aminoactinomycin D (7-AAD; BD Biosciences, San Jose, CA, USA). Isotype-matched antibodies were used as controls. Cells were acquired using a BD LSRFortessa X-20 (BD Biosciences, San Jose, CA, USA), and data were analyzed using FlowJo version 10.10.1 (BD Biosciences).

The gating strategy is shown in Figure 1A. After gating on broad mononuclear cells encompassing the lymphocyte and monocyte regions, CD19^+^ B-cells were identified. Total CD19^+^ B cells were expressed as a percentage of viable mononuclear cells, whereas B-cell subset frequencies were expressed as percentages within the CD19^+^ B-cells. CD21^low^ B-cells were defined as CD21^low^ cells; class-switched B-cells as CD27^+^IgD^−^ cells; plasmablasts as CD27^high^CD38^high^ cells within the class-switched B-cell population; and plasma cells as CD138^+^ cells within the plasmablast population. These phenotypic definitions were based on published classifications of circulating switched B-cells, plasmablasts, and plasma cells (18–20). Gating and subset quantification were performed by an investigator blinded to clinical outcome, and all gates were reviewed by a second investigator.

### 2.4 B-cell receptor repertoire analysis

Paired pretreatment and week-1 immunoglobulin G (IgG)-targeted human immunoglobulin heavy-chain repertoire data were available from PBMCs in four patients with severe irAEs. Paired repertoires from CD21^low^ B-cells, isolated by fluorescence-activated cell sorting within the CD19^+^ B-cell gate, were available from three of these patients. Cell sorting was performed using a FACSAria III (BD Biosciences, San Jose, CA, USA). Total RNA was extracted from PBMCs or sorted CD21^low^ B-cells using TRIzol LS (Thermo Fisher Scientific, Waltham, MA, USA), purified using an RNeasy Mini Kit, and assessed using an Agilent 2200 TapeStation (Agilent Technologies, Santa Clara, CA, USA).

IgG B-cell receptor (BCR) repertoire analysis was performed using unbiased next-generation sequencing developed by Repertoire Genesis, Inc. (Ibaraki, Osaka, Japan), in accordance with previous studies (21, 22). BCR sequences were grouped according to identical IGHV, IGHJ, IGHC, and CDR3 amino-acid sequences. To normalize sequencing depth, repertoires were rarefied to 100,000 reads by random sampling without replacement. Rarefaction was repeated 1,000 times, and mean values across iterations were used for analysis. Shannon diversity, Pielou’s evenness, DE50, and the Gini coefficient were calculated (23, 24).

### 2.5 Statistical analysis

Patient-level week-1 fold changes were compared between patients with and without severe irAEs using exact two-sided Mann-Whitney U tests. P values were not adjusted for multiple comparisons. Statistical analyses and graph generation were performed using GraphPad Prism version 11.0.2 (GraphPad Software). Custom repertoire-analysis scripts were implemented in Python version 3.13.14 using NumPy version 2.5.1 and pandas version 3.0.3. Excel repertoire reports were imported using openpyxl version 3.1.5.

## 3 Results

### 3.1 Fluctuations of activated B-cell subsets after nivolumab plus ipilimumab

To characterize changes in activated B-cell subsets after nivolumab plus ipilimumab, we performed flow-cytometric analyses of CD21^low^ B-cells, class-switched B-cells, plasmablasts, and plasma cells within the CD19^+^ B-cell compartment (Figure 1A). In an initial exploratory analysis, the proportions of these activated B-cell subsets increased as early as 1 week after the first nivolumab plus ipilimumab administration, then declined toward baseline before the next treatment cycle (Figure 1B). Based on this observation, we then analyzed peripheral blood B-cell subsets at pretreatment, week 1, and before cycle 2 in 12 patients who received nivolumab plus ipilimumab to examine the association between these changes and severe irAEs. Extended longitudinal analysis in patients 1 and 3 (Table 1) showed similar transient increases in these B-cell subsets after subsequent administrations of nivolumab plus ipilimumab (Figure 2).

**Table 1.** Patient characteristics, treatment doses, and immune-related adverse events in the nivolumab-plus-ipilimumab cohort.

| Patient ID | Age group, years | Sex | Cancer type | Stage | Nivolumab dose (mg/body) | Ipilimumab dose (mg/kg) | irAE type and grade | Max grade of irAEs | Time to grade ≥3 irAE, days |
| --- | --- | --- | --- | --- | --- | --- | --- | --- | --- |
| Patient 1 | 60-69 | Male | Melanoma | Stage IV | 80 | 3 | Dermatitis G3 / Colitis G1 | 3 | 29 |
| Patient 2 | 30-39 | Male | Renal cell carcinoma | Stage IV | 240 | 1 | Hepatitis G3 | 3 | 21 |
| Patient 3 | 70-79 | Male | Renal cell carcinoma | Stage IV | 240 | 1 | None | 0 |  |
| Patient 4 | 60-69 | Male | Renal cell carcinoma | Stage IV | 240 | 1 | Dermatitis G1 | 1 |  |
| Patient 5 | 70-79 | Female | Melanoma | Stage IV | 80 | 3 | None | 0 |  |
| Patient 6 | 60-69 | Female | Melanoma | Stage IV | 80 | 3 | Myasthenia gravis G3 / Thyroiditis G2 | 3 | 85 |
| Patient 7 | 40-49 | Male | Melanoma | Stage IV | 80 | 3 | Adrenal failure G2 / Thyroiditis G2 | 2 |  |
| Patient 8 | 70-79 | Male | Renal cell carcinoma | Stage IV | 240 | 1 | None | 0 |  |
| Patient 9 | 70-79 | Male | Renal cell carcinoma | Stage IV | 240 | 1 | Thyroiditis G2 | 2 |  |
| Patient 10 | 70-79 | Male | Renal cell carcinoma | Stage IV | 240 | 1 | Nephritis G3 | 3 | 33 |
| Patient 11 | 70-79 | Male | Renal cell carcinoma | Stage IV | 240 | 1 | Adrenal failure G3 / Thyroiditis G2 | 3 | 42 |
| Patient 12 | 50-59 | Male | Renal cell carcinoma | Stage IV | 240 | 1 | Thyroiditis G1 / Dermatitis G1 | 1 |  |

Patient characteristics, treatment regimens, irAEs, highest irAE grade, and time to the first severe irAE are shown. Ages are presented in non-overlapping groups of ten years. Blank cells indicate that no severe irAE occurred during the 6-month observation period. irAE, immune-related adverse event.

**Figure 2.**
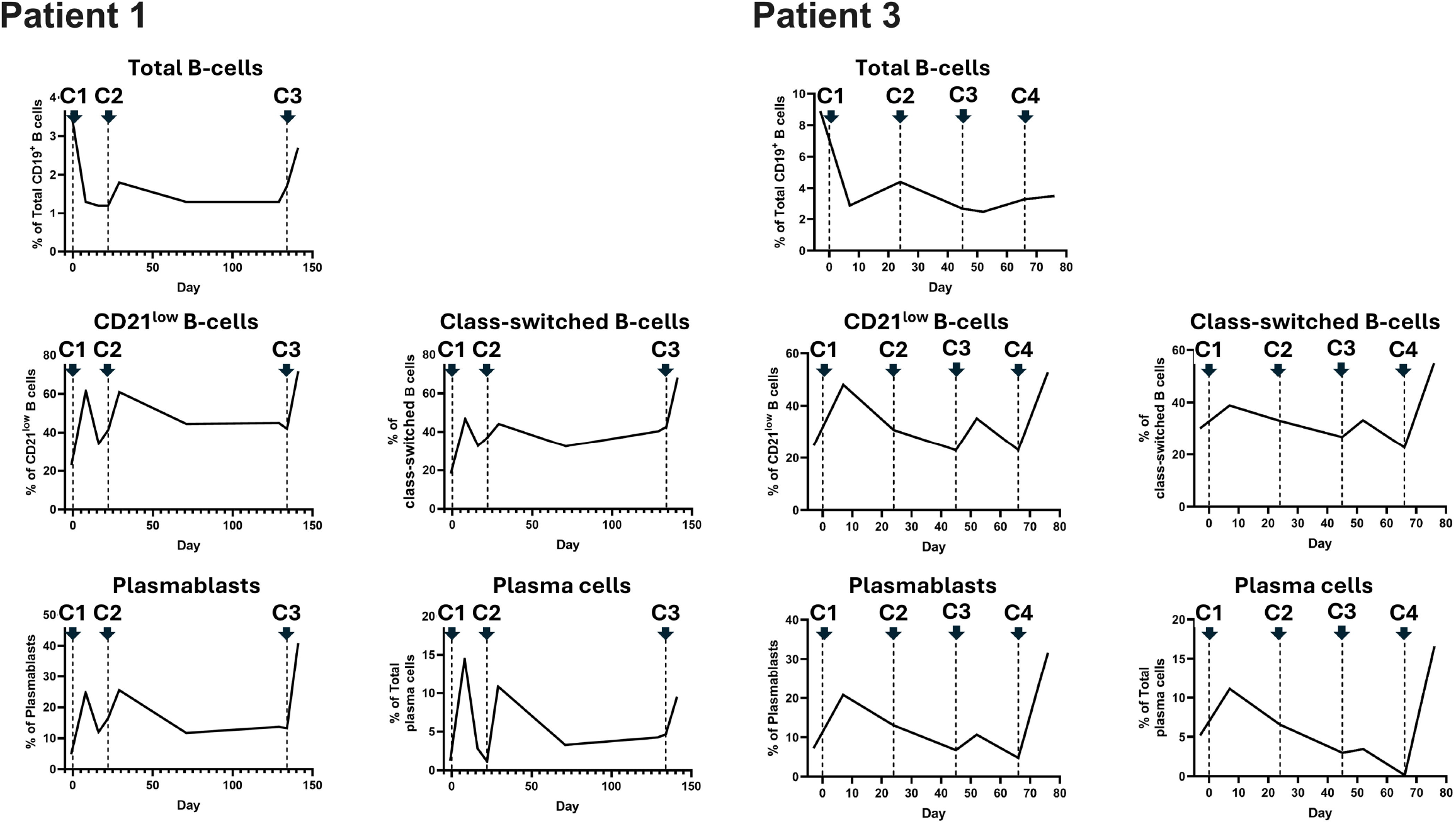
Recurrent changes in activated B-cell populations after repeated nivolumab plus ipilimumab. Serial changes in total CD19^+^ B-cells, CD21^low^ B-cells, CD27^+^IgD^−^ class-switched B-cells, plasmablasts, and CD19^+^CD138^+^ plasma cells are shown for patients 1 and 3, who had additional samples. Total CD19^+^ B-cells are expressed as a percentage of viable mononuclear cells, whereas B-cell subset frequencies are expressed as percentages of CD19^+^ B cells. Downward arrows and vertical dashed lines indicate administration of nivolumab plus ipilimumab on Day 1 of each treatment cycle. C, treatment cycle; PBMC, peripheral blood mononuclear cell. Alt text: Serial line plots show total CD19^+^ B-cells and four activated B-cell subsets across repeated nivolumab-plus-ipilimumab treatment cycles in two patients. These subsets show recurrent post-treatment increases of varying magnitude after successive treatment cycles.

### 3.2 Association between first-week B-cell changes and severe irAEs

The four activated B-cell subsets generally increased at week 1 and declined before cycle 2 (Figure 3A). During the 6-month observation period, 5 of 12 patients developed severe irAEs (Table 1).

**Figure 3.**
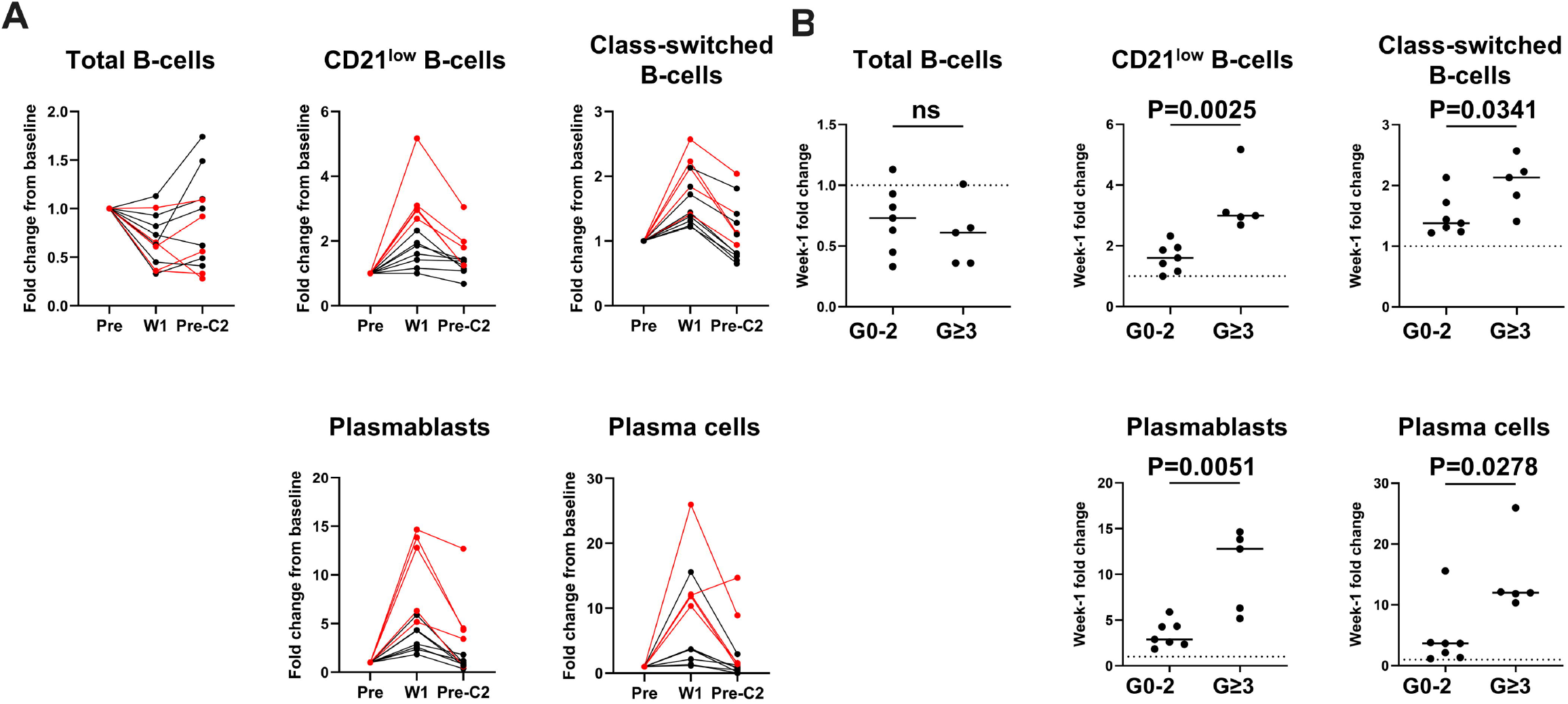
Early B-cell changes after nivolumab plus ipilimumab and severe irAEs. (A) Patient-level fold changes from pretreatment at week 1 and before cycle 2. Red lines indicate patients who developed severe irAEs; black lines indicate patients who did not. (B) Week-1 fold changes according to subsequent severe irAE status. Each point represents one patient: horizontal lines indicate medians. P values were calculated using exact two-sided Mann-Whitney U tests. The dotted horizontal line indicates no change from pretreatment (fold change = 1). irAE, immune-related adverse event; Pre, Pretreatment; W1, 1 week after the first dose; Pre-C2, before cycle 2; G, Grade; ns, not significant. Alt text: Patient-level plots show changes in total and activated B-cell populations from pretreatment to week 1 and before cycle 2 after nivolumab plus ipilimumab. Patients who subsequently developed severe irAEs show greater week-1 increases in four activated B-cell subsets than those without severe events, whereas total CD19+ B-cell changes do not differ between groups.

Week-1 fold changes in these subsets were significantly greater in patients with severe irAEs than in those without (Figure 3A and B). Week-1 fold changes in total CD19^+^ B-cells did not differ between groups.

We also examined five patients who received anti-PD-1 monotherapy. Their clinical characteristics and irAE profiles are summarized in Supplementary Table 1. This exploratory group showed no consistent pattern of changes at week 1 across the four activated B-cell populations (Supplementary Figure 1). In one patient in this exploratory group who developed a severe irAE, all four activated B-cell subsets increased at week 1 compared with pretreatment.

### 3.3 Changes in IgG BCR repertoires after nivolumab plus ipilimumab

Among the five patients who developed severe irAEs, paired pretreatment and week-1 IgG BCR repertoires derived from PBMCs were available for four patients. Three of these four patients also had paired pretreatment and week-1 repertoires from sorted CD21^low^ B-cells. Diversity metrics showed an overall tendency to increase from pretreatment to week 1 (Figure 4A). In PBMC-derived repertoires, DE50 increased in all four pairs, and both Shannon diversity and Pielou’s evenness increased in three of the four pairs. In sorted CD21^low^ B-cell repertoires, all three metrics increased in all three pairs (Figure 4A). To further characterize the clonotype abundance distribution, we assessed the Gini coefficient. The Gini coefficient decreased from pretreatment to week 1 in all four PBMC-derived repertoire pairs and all three sorted CD21^low^ B-cell repertoire pairs, indicating reduced clonotype concentration (Figure 4B). This finding was consistent with the observed increase in repertoire diversity. Unrarefied diversity and Gini coefficient analyses showed the same overall pattern (Supplementary Figure 2).

**Figure 4.**
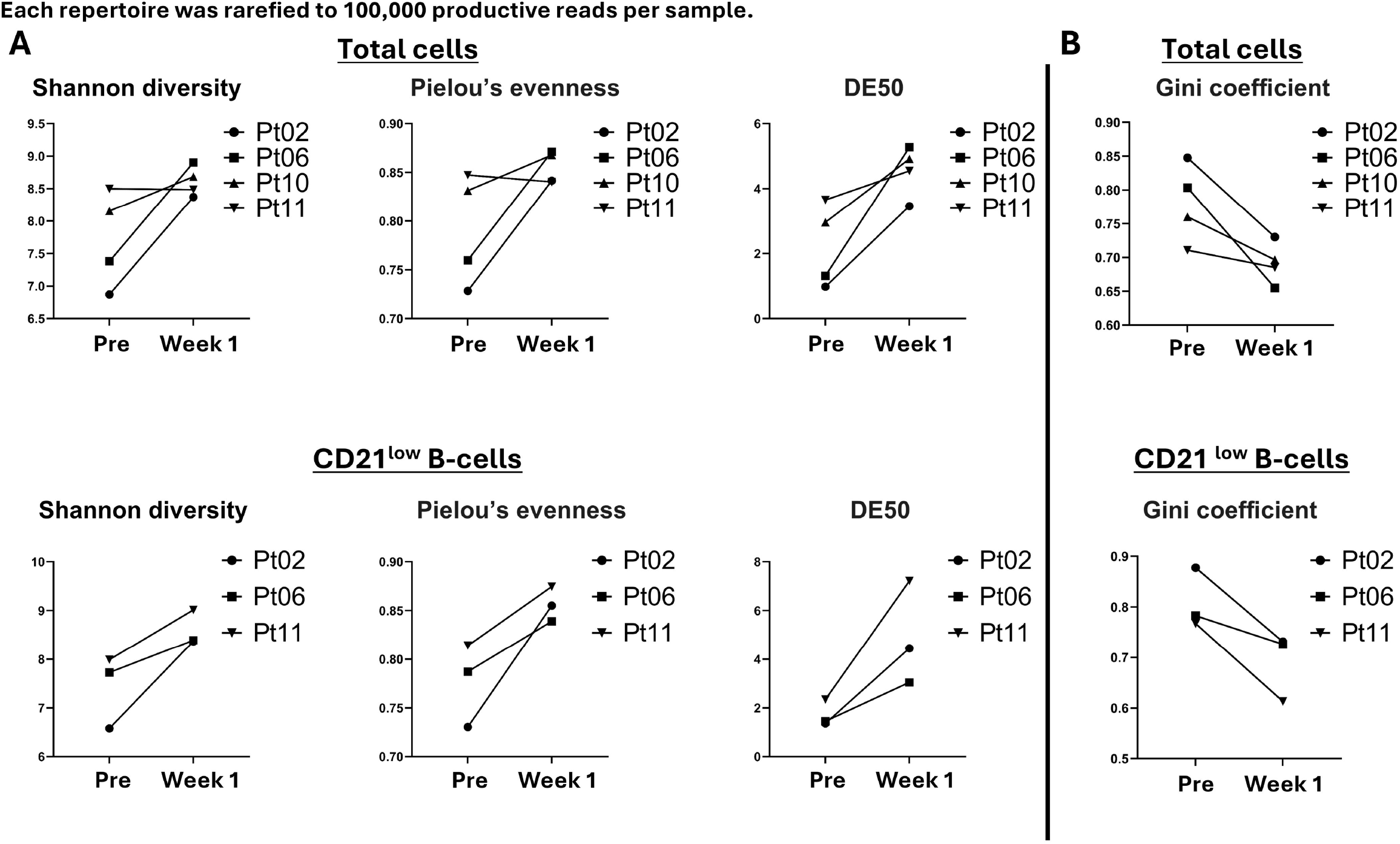
Paired IgG B-cell receptor repertoire diversity and clonotype abundance distribution. Immunoglobulin G (IgG) B-cell receptor (BCR) repertoire metrics were calculated after rarefaction to 100,000 productive reads per sample. (A) Shannon diversity, Pielou’s evenness, and DE50 before treatment and at week 1 in PBMC-derived repertoires (n = 4) and sorted CD21^low^ B-cell repertoires (n = 3). Lines connect paired samples from the same patient. (B) Gini coefficients before treatment and at week 1 in PBMC-derived repertoires (n = 4) and sorted CD21^low^ B-cell repertoires (n = 3). Lower Gini coefficients indicate a more even distribution of reads across clonotypes. Alt text: Paired plots compare IgG B-cell receptor repertoires before treatment and at week 1. In PBMC-derived repertoires, DE50 increases in all four patients and Shannon diversity and Pielou’s evenness increase in three of four patients; all three diversity measures increase in all three CD21^low^ B-cell repertoires. The Gini coefficient decreases in all paired repertoires, indicating a more even overall distribution of reads across clonotypes.

### 3.4 Anti-acetylcholine receptor antibody elevation before the onset of myasthenia gravis symptoms

Patient 6 was treated with nivolumab plus ipilimumab, and subsequently developed ICI-related myasthenia gravis (MG). The anti-acetylcholine receptor (AChR) antibody concentration was 45.0 nmol/L on Day 81, and MG was diagnosed by a neurologist on Day 85 (Figure 5). After the diagnosis, archived serum samples collected before and after the first treatment were retrospectively tested for anti-AChR antibodies. Anti-AChR antibody was below the reporting threshold before treatment (<0.1 nmol/L), but had already exceeded the laboratory reference range (≤0.2 nmol/L) by Day 11 (1.8 nmol/L). It continued to increase thereafter, indicating that an AChR-directed antibody response had emerged early after treatment initiation, before the onset of MG symptoms.

**Figure 5.**
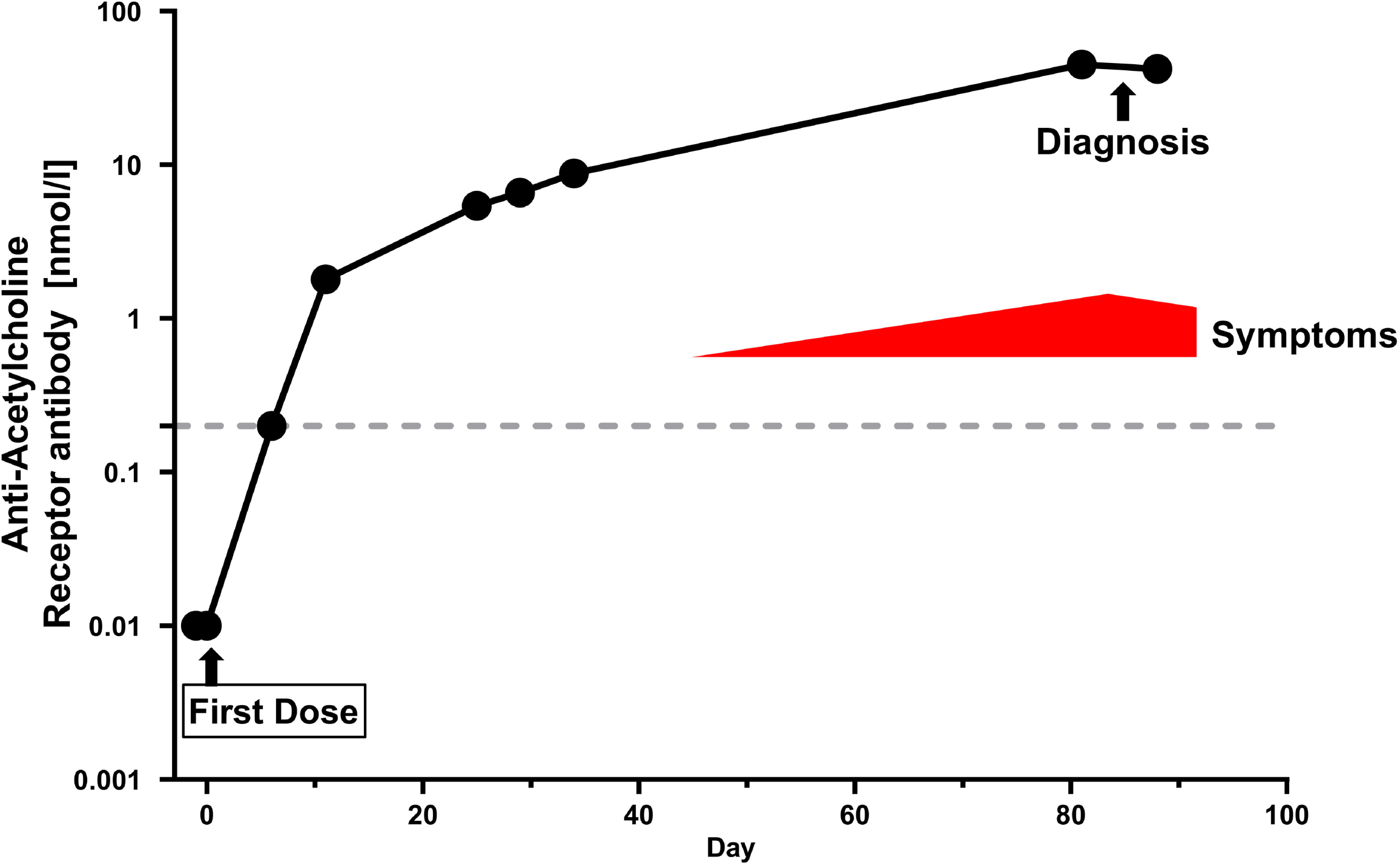
Integrated clinical, serologic, and neurologic timeline of the myasthenia gravis case. Serial anti-acetylcholine receptor (AChR) antibody concentrations and the clinical course of immune checkpoint inhibitor-related myasthenia gravis (MG) in patient 6 are shown. After MG was diagnosed, archived serum samples obtained before and after nivolumab-plus-ipilimumab initiation were tested retrospectively by radioimmunoassay at LSI Medience Corporation (Tokyo, Japan). The dashed horizontal line indicates the upper limit of the reference range (0.2 nmol/L). Values below the laboratory reporting threshold (<0.1 nmol/L) were plotted at 0.01 nmol/L solely for visualization on the logarithmic axis. The red area indicates the period of neurologic symptoms. The arrow at Day 0 indicates the first administration of nivolumab plus ipilimumab, and the black arrow indicates the diagnosis of MG. Alt text: A logarithmic time-course plot shows anti-AChR antibody concentrations after nivolumab plus ipilimumab in a patient who developed myasthenia gravis. The antibody level is below the reporting threshold before treatment, exceeds the laboratory reference range by Day 11, and continues to increase before the onset of neurologic symptoms and the diagnosis of myasthenia gravis on Day 85.

## 4 Discussion

In this study, nivolumab plus ipilimumab induced marked changes in circulating activated B-cell populations as early as 1 week after treatment initiation. Increases in CD21^low^ B-cells, class-switched B-cells, plasmablasts, and plasma cells at week 1 were greater in patients who later developed severe irAEs. These findings suggest that early changes in the activation profile of the peripheral B-cell compartment were associated with the subsequent development of severe irAEs. IgG BCR repertoire diversity also increased in patients who developed severe irAEs, supporting the presence of an early B-cell immune response. In addition, anti-AChR antibodies were already elevated by Day 11 in the patient who later developed ICI-related myasthenia gravis, supporting the biological relevance of early B-cell activation to the subsequent development of irAEs.

Some irAEs have been suggested to share features with autoimmune diseases involving antigen-antibody interactions (11). On this basis, we hypothesized that changes across antigen-experienced and antibody-secreting B-cell subsets might be associated with irAEs, and therefore performed a detailed analysis of these subsets. CD21^low^ B-cells comprise a heterogeneous population that includes atypical memory B-cells and is expanded in conditions associated with chronic immune stimulation and autoimmunity (25, 26). Das et al. reported that increases in CD21^low^ B-cells after combined PD-1 and CTLA-4 blockade were associated with irAEs (27). The present study extends this observation by evaluating multiple B-cell subsets at week 1, including class-switched B-cells, plasmablasts, and plasma cells. These B-cell subsets may contribute to immune dysregulation associated with irAEs through antigen presentation, cytokine production, and autoantibody production (11, 28).

To further characterize early B-cell activation, we analyzed IgG BCR repertoires in patients who subsequently developed severe irAEs. Repertoire diversity generally increased from pretreatment to week 1 in PBMC-derived repertoires and consistently increased in sorted CD21^low^ B-cell repertoires. The Gini coefficient, which reflects the extent to which repertoire reads are concentrated, also decreased in all paired repertoires. Taken together, the increase in repertoire diversity and the reduced tendency of reads to concentrate in a limited number of clonotypes suggest that the IgG BCR repertoire became broader and more evenly distributed by week 1. Such early repertoire changes may reflect a broader potential range of B-cell antigen recognition, possibly including self-antigens (10). Collectively, the repertoire data support the presence of an early B-cell immune response in patients who subsequently developed severe irAEs.

Although irAEs often do not become clinically apparent until 2–3 months after ICI initiation, it is noteworthy that B-cell changes as early as 1 week after the first dose were associated with their subsequent development (14, 15). From a general immunological perspective, humoral immune responses can emerge approximately 1 week after infection or vaccination, and our previous BCR repertoire analyses following SARS-CoV-2 vaccination demonstrated detectable B-cell responses around 7 days after antigen exposure (21, 22). Importantly, in the patient who subsequently developed ICI-related myasthenia gravis, anti-acetylcholine receptor antibody was already increased by Day 11. Because detectable antibody production requires preceding B-cell activation and differentiation into antibody-secreting cells, this temporal course supports the biological plausibility that B-cell activation was already underway around the first week after treatment initiation and may have been involved in the subsequent development of irAEs.

This exploratory study has several limitations. First, the sample size was small and the study was conducted at a single institution, which limits the statistical power and generalizability of the findings. Second, although the observed B-cell changes were associated with subsequent severe irAEs, the study did not establish that they were specific to the immune processes underlying the irAEs; they may also have reflected general pharmacodynamic immune activation induced by nivolumab plus ipilimumab. Third, BCR repertoire analyses were limited to patients with severe irAEs, and comparator repertoire data from patients without such events were unavailable. These findings therefore require validation in a larger prospective cohort.

In conclusion, early activation of peripheral B-cell subsets at 1 week after nivolumab plus ipilimumab was associated with the subsequent development of severe irAEs.

## 5 Data Availability Statement

All data produced in the present study are available upon reasonable request to the authors.

## 6 Ethics Statement

### Ethics statement

This study was conducted in accordance with the principles of the Declaration of Helsinki.

### Participant consent statement

All participants provided written informed consent for this research.

### Ethics approval

The study protocol was approved by Kobe University Hospital Ethics Committee (approval numbers B2056704 and 1481).

## 7 Author Contributions

HT: Writing – original draft, Writing – review & editing, Data curation, Formal analysis, Software, Funding acquisition, Conceptualization. YF: Conceptualization, Data curation, Formal analysis, Investigation, Project administration, Resources, Supervision, Writing – original draft, Writing – review & editing. TK: Resources, Writing – review & editing. TS: Writing – review & editing. EF: Writing – review & editing. JML: Writing – review & editing. KK: Writing – review & editing. YI: Writing – review & editing. KY: Writing – review & editing. SK: Writing – review & editing. YN: Writing – review & editing. NK: Writing – review & editing. HM: Conceptualization, Funding acquisition, Supervision, Writing – review & editing.

All authors contributed to the article and approved the submitted version.

## 8 Funding

This work was supported by JSPS KAKENHI Grant Number JP23H02766.

## 9 Acknowledgments

During the final preparation of this manuscript, the authors used ChatGPT (OpenAI, San Francisco, CA, USA; model: GPT-5.6 Sol; version accessed in August 2026; https://chatgpt.com/) solely for English-language proofreading and stylistic editing. All AI-assisted revisions were reviewed and approved by the authors, who take full responsibility for the accuracy and content of the final manuscript.

## 10 Conflict of Interest

T.K. has received speaker fees from Merck Biopharma, Novartis Pharma, Daiichi Sankyo, Eisai, and MSD, and research funding to the institution from AstraZeneca. T.S. is an employee of KAICO Ltd. The other authors declare no potential conflicts of interest. K.Y. has received research grants and honoraria from Pfizer. Y.N. has received speaking and lecture fees from Ono Pharmaceutical, Bristol Myers Squibb, MSD, Chugai Pharmaceutical, Takeda Pharmaceutical, Taiho Pharmaceutical, Eli Lilly, and Astellas Pharma. N.K. has received lecture fees from Ono Pharmaceutical, Bristol Myers Squibb, MSD, Merck Biopharma, Chugai Pharmaceutical, Bayer, Eisai, Daiichi Sankyo, and Novartis Pharma, and research funding from Ono Pharmaceutical, AstraZeneca, Rakuten Medical, Adlai Nortye, Boehringer Ingelheim, GSK, and AbbVie. H.M. has received lecture fees from Daiichi Sankyo and Chugai Pharmaceutical, research funding from AbbVie, Chugai Pharmaceutical, GSK, Kyowa Kirin, and Taiho Pharmaceutical, and contributions from Chugai Pharmaceutical.

The remaining authors declare that the research was conducted in the absence of any commercial or financial relationships that could be construed as a potential conflict of interest.

## 11 Permission to Reproduce Material from Other Sources

N/A

## Notes

### Competing Interest Statement

T.K. received speaker fees from Merck Biopharma, Novartis Pharma, Daiichi Sankyo, Eisai, and MSD, and institutional research funding from AstraZeneca. T.S. is an employee of KAICO Ltd. K.Y. received research grants and honoraria from Pfizer. Y.N. received speaking and lecture fees from Ono Pharmaceutical, Bristol Myers Squibb, MSD, Chugai Pharmaceutical, Takeda Pharmaceutical, Taiho Pharmaceutical, Eli Lilly, and Astellas Pharma. N.K. received lecture fees from Ono Pharmaceutical, Bristol Myers Squibb, MSD, Merck Biopharma, Chugai Pharmaceutical, Bayer, Eisai, Daiichi Sankyo,
and Novartis, and research funding from Ono Pharmaceutical, AstraZeneca, Rakuten Medical, Adlai Nortye, Boehringer Ingelheim, GSK, and AbbVie. H.M. received lecture fees from Daiichi Sankyo and Chugai Pharmaceutical, research funding from AbbVie, Chugai Pharmaceutical, GSK, Kyowa Kirin, and Taiho Pharmaceutical, and contributions from Chugai Pharmaceutical.
The remaining authors declare no competing interests.

### Author Declarations

The Kobe University Hospital Ethics Committee of Kobe University Hospital gave ethical approval for this work (approval numbers B2056704 and 1481).

