## Supplementary Table 1 and Supplementary Figures 1 and 2 for "First-week expansion of activated peripheral B-cell subsets is associated with grade 3 or higher immune-related adverse events after nivolumab plus ipilimumab"

Supplementary Material

**Supplementary Table 1. Clinical characteristics and immune-related adverse events in patients treated with anti-PD-1 monotherapy**

| **Patient ID** | **Age group, years** | **Sex** | **Cancer type** | **Stage** | **Nivolumab dose (mg/body)** | **Pembrolizumab dose (mg/body)** | **irAE type and grade** | **Max grade of irAEs** | **Time to grade ≥3 irAE, days** |
| --- | --- | --- | --- | --- | --- | --- | --- | --- | --- |
| Patient 13 | 30-39 | Male | Eyelid cancer | Stage IV | 240 |  | Dermatitis G1 | 1 |  |
| Patient 14 | 70-79 | Female | Mucosal melanoma | Stage IV | 240 |  | Hepatitis G3 | 3 | 18 |
| Patient 15 | 60-69 | Male | Gastric cancer | Stage IV | 240 |  | None | 0 |  |
| Patient 16 | 70-79 | Male | Mucosal melanoma | Stage IV |  | 200 | None | 0 |  |
| Patient 17 | 70-79 | Female | Tongue cancer | Stage IV | 240 |  | None | 0 |  |

Clinical characteristics, anti-PD-1 agents, treatment doses, irAEs, highest irAE grade, and time to the first grade 3 or higher irAE are shown for the exploratory monotherapy group. Ages are presented in non-overlapping groups of ten years. Blank cells indicate that drug was not administered or that no grade 3 or higher irAE occurred. irAE, immune-related adverse event; PD-1, programmed cell death protein 1.

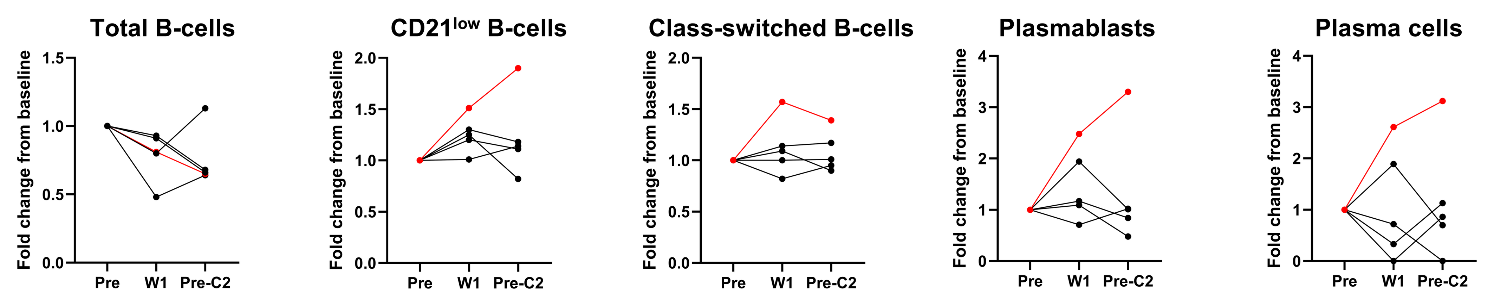

**Supplementary Figure 1. Serial B-cell changes in the anti-PD-1 monotherapy group** Patient-level fold changes from pretreatment at week 1 and before cycle 2 are shown for total CD19^+^ B-cells, CD21^low^ B-cells, CD27^+^IgD^−^ class-switched B-cells, plasmablasts, and CD19^+^CD138^+^ plasma cells in five patients treated with anti-PD-1 monotherapy. The red line indicates the patient who developed a grade 3 irAE; black lines indicate the other four patients. irAE, immune-related adverse event; PD-1, programmed cell death protein 1; Pre, Pretreatment; W1, 1 week after the first dose; Pre-C2, before cycle 2.

Alt text: Patient-level plots show changes in five peripheral B-cell populations from pretreatment to week 1 and before cycle 2 in five patients receiving anti-PD-1 monotherapy. No consistent week-1 pattern is observed across the group; however, the single patient who developed a grade 3 immune-related adverse event shows week-1 increases in CD21^low^ B-cells, class-switched B-cells, plasmablasts, and plasma cells.

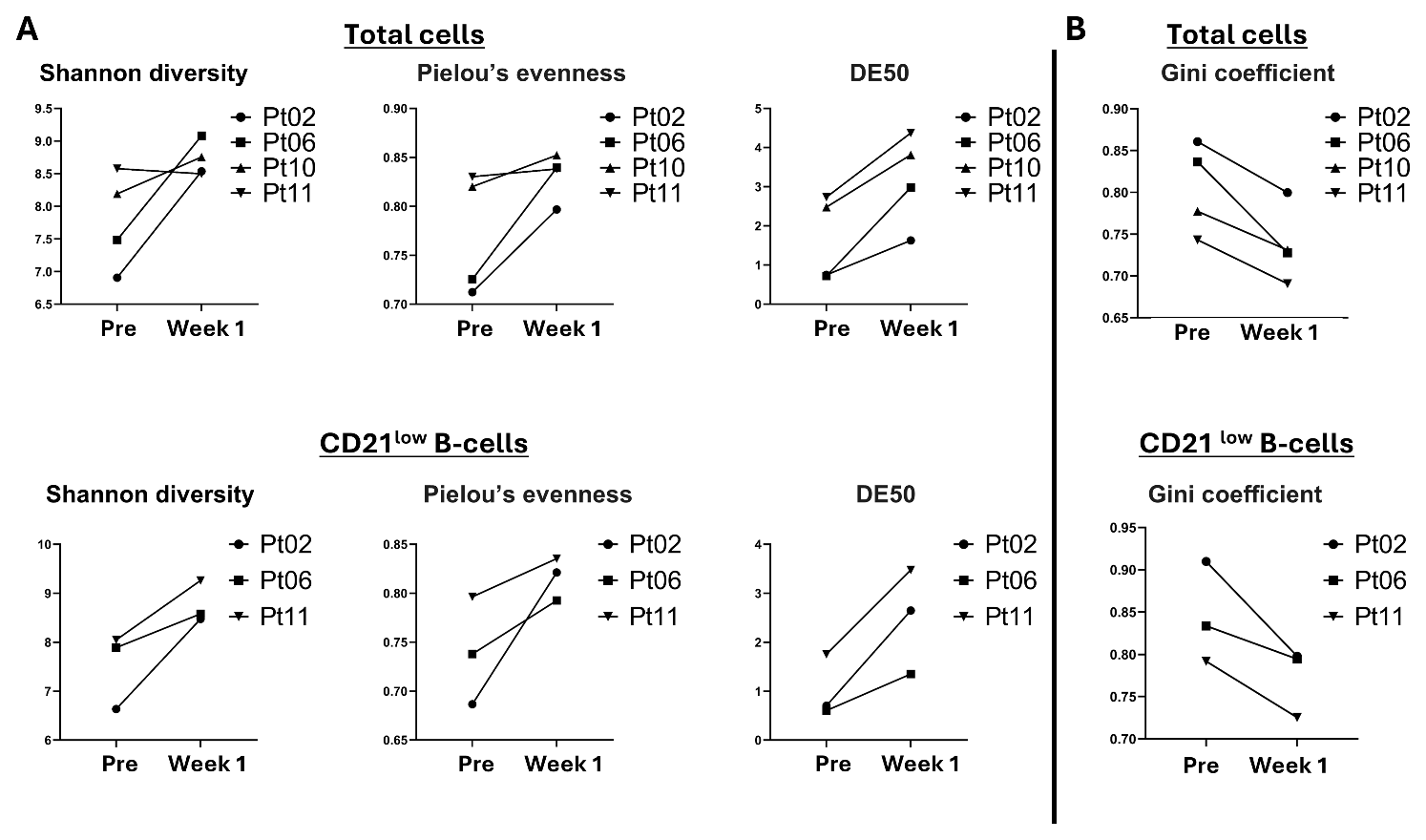

**Supplementary Figure 2. Paired IgG B-cell receptor repertoire diversity and clonotype abundance distribution without rarefaction** Sensitivity analyses using unrarefied IgG B-cell receptor (BCR) repertoire data are shown. (A) Shannon diversity, Pielou's evenness, and DE50 before treatment and at week 1 in PBMC-derived repertoires (n = 4) and sorted CD21^low^ B-cell repertoires (n = 3). Lines connect paired samples from the same patient. (B) Gini coefficients before treatment and at week 1 in PBMC-derived repertoires (n = 4) and sorted CD21^low^ B-cell repertoires (n = 3). PBMC, peripheral blood mononuclear cell.

Alt text: Paired line plots compare unrarefied immunoglobulin G B-cell receptor repertoires before treatment and at week 1. Panel A shows generally increased Shannon diversity, Pielou’s evenness, and DE50 in PBMC-derived and sorted CD21low B-cell repertoires. Panel B shows decreased Gini coefficients in all paired repertoires, indicating a more even distribution of reads across clonotypes.
